# *Can you hear me now*? Clinical applications of audio recordings

**DOI:** 10.1101/2022.02.07.22270598

**Authors:** Anish Kumar, Theo Jaquenoud, Jacqueline Helcer Becker, Dayeon Cho, Monica Rivera Mindt, Alex Federman, Gaurav Pandey

## Abstract

Audio and speech have several implicit characteristics that have the potential for the identification and quantification of clinical disorders. This PRISMA-guided review is designed to provide an overview of the landscape of automated clinical audio processing to build data-driven predictive models and infer phenotypes of a variety of neuropsychiatric, cardiac, respiratory and other disorders. We detail the important components of this processing workflow, specifically data acquisition and processing, algorithms used and their customization for clinical applications, commonly used tools and software, and benchmarking and evaluation methodologies. Finally, we discuss important open challenges for the field, and potential strategies for addressing them.

## Introduction

Audio has long served as a rich source of information for clinicians to evaluate the health of their patients.^1,2^ Routine physical exams involve auscultation or listening to various sounds (respiratory, cardiac, gastrointestinal, etc.), which indicate the health of various organs and physiologic systems.^2^ Many clinicians also consider patients’ speech patterns to assess mood, cognition, and other neurological functions.^1,3–9^

Audio and speech also have implicit characteristics that human experts are unable to hear and quantify.^10^ However, with improvements in computational power and machine learning algorithms,^11,12^ data scientists have recently been able to unlock novel dimensions of audio and provide clinicians with more information to support decision-making.^10^ We present an overview of this work on developing novel biomarkers, predictive models and other data-driven inferences from clinical audio for a variety of disorders. We detail various components of the audio processing workflow (**Figure 1**), beginning with a description of the considerations and tools for collecting and processing clinical audio data. We then present an overview of statistical, traditional machine learning,^11^ and deep learning^13^ methods that have been leveraged for the analysis of clinical audio for diagnostic and prognostic purposes. We end with a discussion of the challenges and opportunities for automated analyses of clinical audio data.

**Figure 1:**
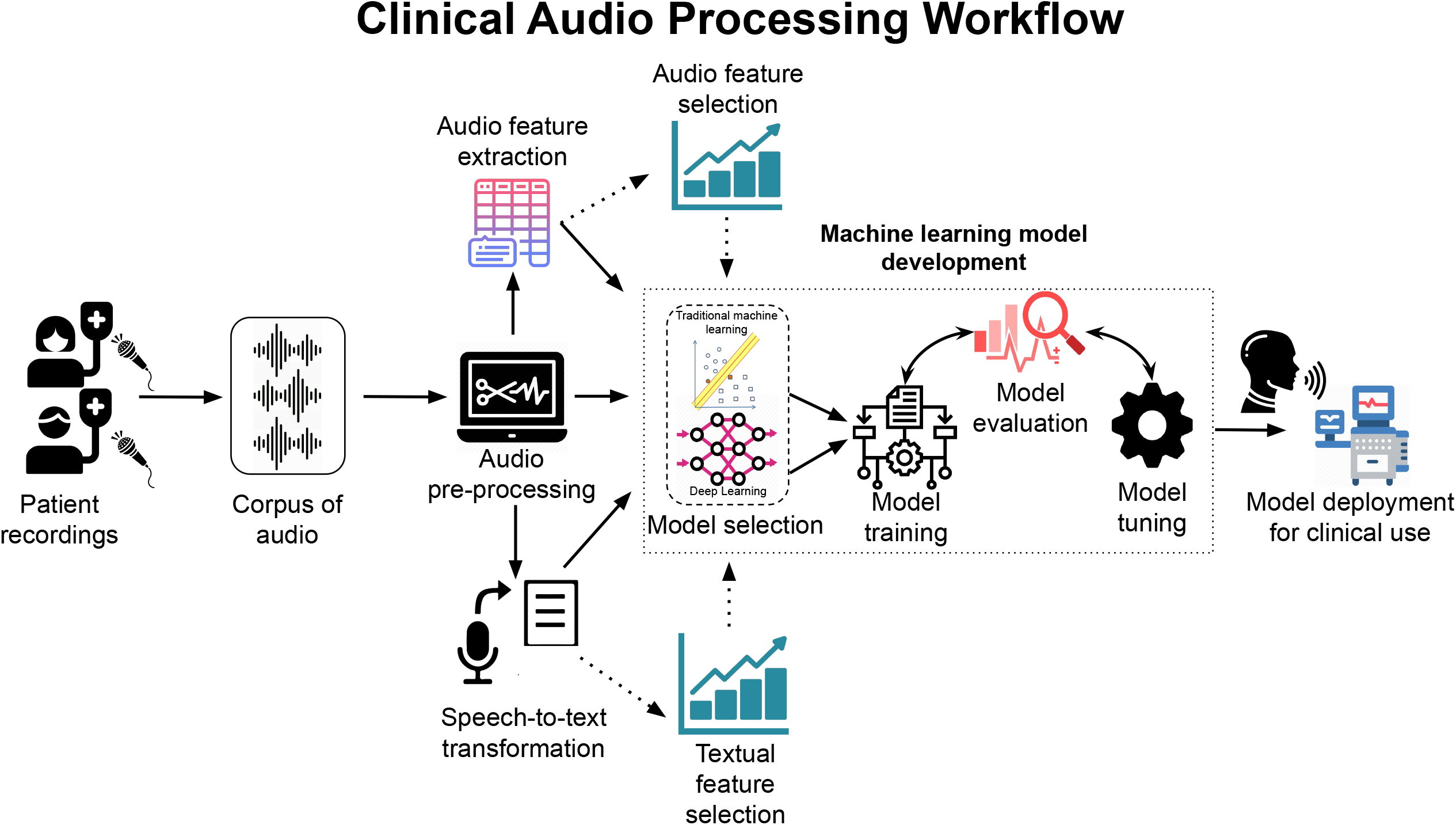
A general workflow for audio processing for clinical use. The process begins with acquisition and compilation of audio recordings from patients. The audio almost always has to be pre-processed to prepare it for downstream modeling. In a feature-based approach, features are extracted from the audio and analyzed. If working with patient speech, an optional step is to perform a speech-to-text transcription and extract textual or linguistic features. Deep learning approaches are often able to work directly with the native audio signal, without a need to explicitly extract features. In the most common applications of clinical audio, a machine learning model is then trained, evaluated and tuned from either the extracted features or the native audio signal. The model can then be integrated into an application and deployed for clinical use. Attributions: Icons in this image were taken from open-source websites, namely www.flaticon.com, www.iconfinder.com, www.clipart-library.com, www.nicepng.com, www.typiccor.com, www.kissclipart.com, www.creazilla.com, www.pngall.com and www.pinclipart.com.

## Methods

Our literature search was conducted using the Preferred Reporting Items for Systematic Reviews and Meta-Analyses (PRISMA) methodology^14^ on the PubMed, Web of Science, and Google Scholar databases using the following keywords: “clinical”, “speech”, “automatic analysis”, “health”, “computational”, “machine learning”, “deep learning” and “audio”. Given the rapid developments in clinical audio processing, we focused on research published after 2010 to emphasize the most recent trends in this field. Furthermore, studies that involved analyses of text generated from speech were included only if they were done in conjunction with an analysis of the native audio. For details of studies from before 2010 and/or focusing on clinical speech, we recommend other excellent reviews. ^1,3–5,9^

The PRISMA flow chart in **Figure 2** details the search results and inclusion/exclusion selection criteria. From the initial 1812 records retrieved from the databases, 561 duplicates were automatically removed, and a further 880 were removed after manual screening of titles and abstracts based on specific inclusion/exclusion criteria, leaving 371 in-scope articles. Studies were included if they explicitly used audio or audio-derived features as clinical biomarkers or were reviews of relevant clinical uses of audio signals; studies that did not use audio signals or used them only for obtaining textual transcription, as well as those that considered ultrasound or echocardiogram only for imaging, were excluded. Among these in-scope articles, this review discusses the 69 most recent and least redundant representative papers on clinical audio processing. These studies were considered representative if they proposed a novel solution or a solution to a new problem, or if they published a new clinical audio dataset. Studies that offered redundant methodologies were excluded in favor of newer or more frequently cited papers. The screening and reviewing of the articles was principally conducted by the first authors.

**Figure 2:**
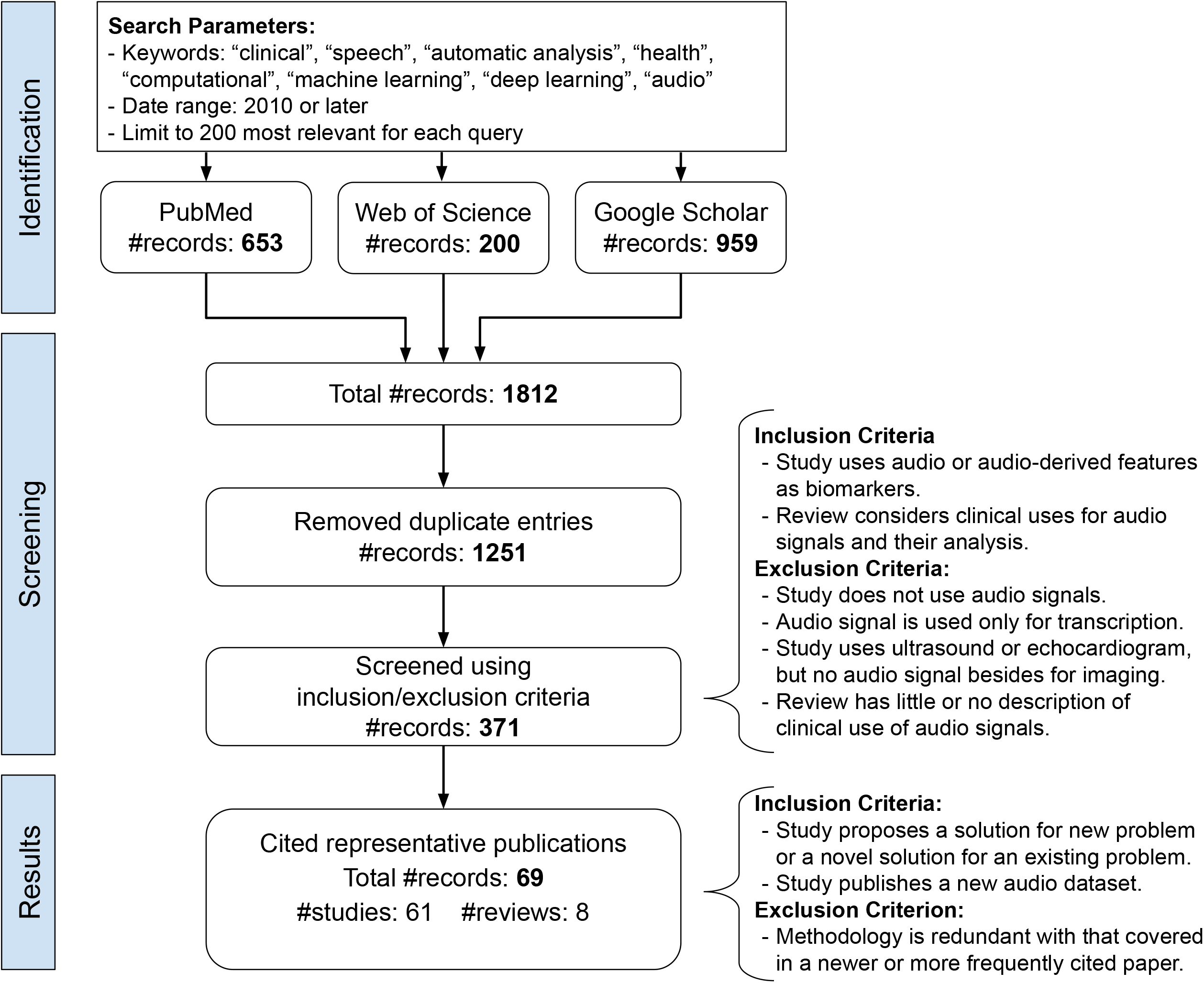
A flowchart detailing the process used to conduct the literature search following PRISMA guidelines. First, records published after 2010 were identified from PubMed, Web of Science, and Google Scholar using the listed keywords. Duplicates were automatically removed. Through manual screening, records are retained/removed according to the specified inclusion/exclusion criteria. The main goal was to only include articles that used or described the use of audio in a clinical setting for purposes other than medical imaging or obtaining transcriptions and performing textual analysis. Of the remaining records, a representative subset were included in this review to accurately portray the scope of clinical applications of audio signals.

### Preliminaries of audio data

Effective data collection and pre-processing are crucial steps in the clinical audio processing workflow (**Figure 1**). Below, we describe several aspects of these tasks.

#### Collection of audio data

A variety of devices were used to record audio in the studies reviewed, ranging from studio-quality microphones to wearable lavaliers, lapel microphones and smartphones. The choice of recording device and its placement relative to the participant may significantly affect data quality.^15^ However, few studies have reported on optimal microphone placement configurations. Some studies suggested using wearable microphones, such as lavaliers, since they are less susceptible to changes in proximity, orientation and posture of the person being recorded.^5^

Other technical factors that can also affect the quality of the recording include the range of frequencies that a microphone can capture. For speech, the recommended upper limit of this range is at least 10 kilohertz (kHz).^15,16^ The choice of sample rate (number of times a data point is recorded per second) and bit depth (number of binary digits used to represent each point) can also affect audio quality. Modern systems generally record at a sample rate of 44.1 kHz and a bit depth of 16 or 24, although some applications require a lower sample rate to improve computational efficiency.^15^

#### Pre-processing audio data

Denoising is the most basic pre-processing of audio data.^17^ While there are sophisticated solutions to this problem,^18^ it is often sufficient to filter out frequencies below 60 Hz, which removes most of the noise induced by common electronic devices.^15^

Normalization is the process of making audio signals from multiple sources, e.g., patients, compatible with each other.^16^ A popular normalization method is min-max scaling, which linearly scales all data from a single source to a given range, typically 0 to 1.^19–21^ Normalization can be performed on the raw audio signal, or more commonly, on features derived from it (next subsection).

When recording a conversation between multiple individuals, e.g., a patient and their physician, it may be necessary to separate the audio signal of each speaker, a task referred to as diarization.^22^ Another useful pre-processing operation is forced alignment, which aligns the audio signals with the transcribed text of the recording.^23^

**Table 1** lists several commonly used tools for audio pre-processing.

**Table 1:**
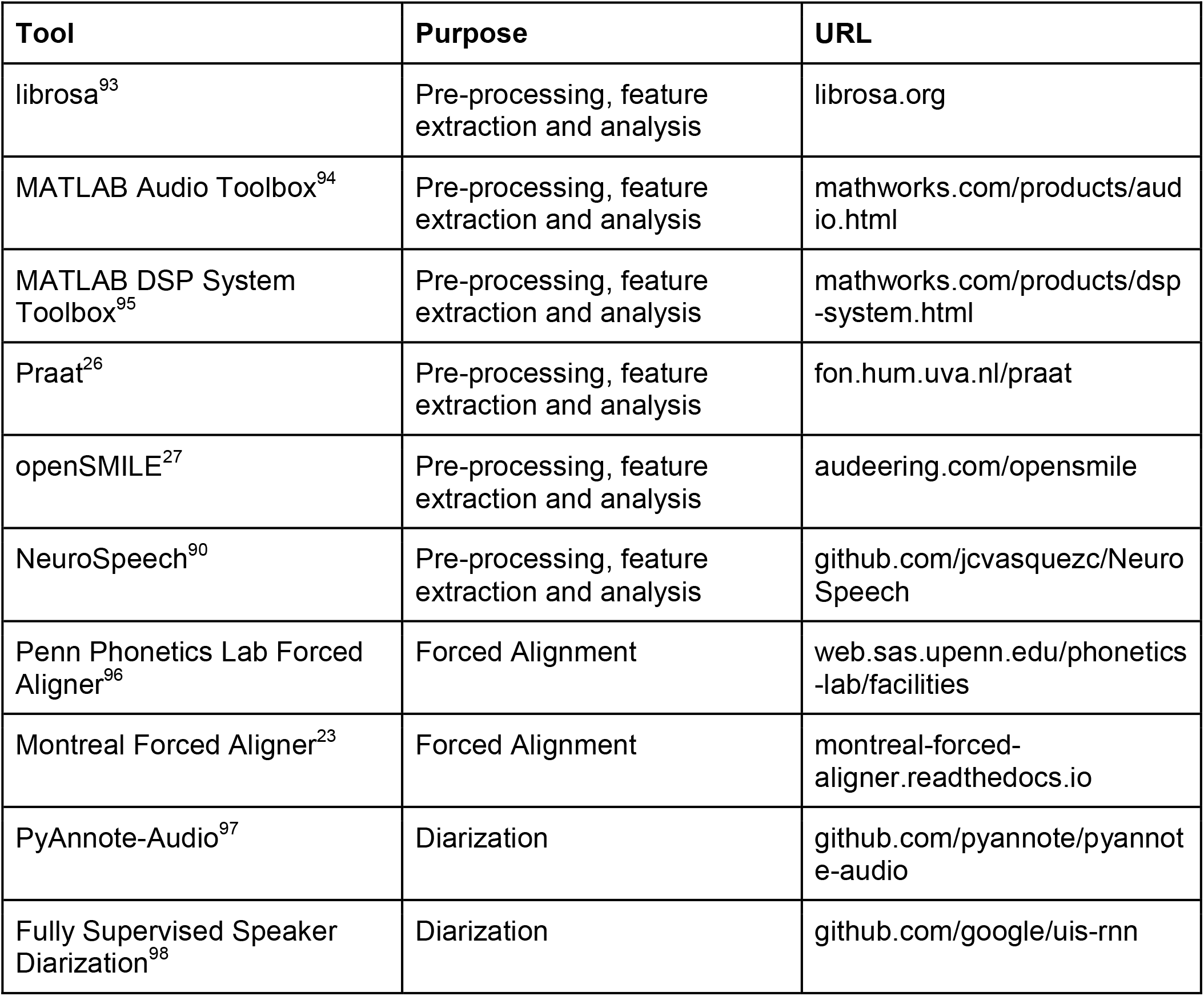
Widely used audio pre-processing and feature extraction tools.

| Tool | Purpose | URL |
| --- | --- | --- |
| librosa<sup>93</sup> | Pre-processing, feature extraction and analysis | librosa.org |
| MATLAB Audio Toolbox<sup>94</sup> | Pre-processing, feature extraction and analysis | mathworks.com/products/audio.html |
| MATLAB DSP System Toolbox<sup>95</sup> | Pre-processing, feature extraction and analysis | mathworks.com/products/dsp-system.html |
| Praat<sup>26</sup> | Pre-processing, feature extraction and analysis | fon.hum.uva.nl/praat |
| openSMILE<sup>27</sup> | Pre-processing, feature extraction and analysis | audeering.com/opensmile |
| NeuroSpeech<sup>90</sup> | Pre-processing, feature extraction and analysis | github.com/jcvasquezc/NeuroSpeech |
| Penn Phonetics Lab Forced Aligner<sup>96</sup> | Forced Alignment | web.sas.upenn.edu/phonetics-lab/facilities |
| Montreal Forced Aligner<sup>23</sup> | Forced Alignment | montreal-forced-aligner.readthedocs.io |
| PyAnnote-Audio<sup>97</sup> | Diarization | github.com/pyannote/pyannote-audio |
| Fully Supervised Speaker Diarization<sup>98</sup> | Diarization | github.com/google/uis-rnn |

#### Extraction of features from audio

In data analysis, a *feature* is defined as a quantitative variable that can be used to describe a data entity.^11^ In the case of (pre-processed) audio, a variety of features can be derived, or *engineered*, as described below.

Audio is natively represented as unstructured waveforms that correspond to variations in sound pressure over time. The features derived from this representation can be broadly related to time, pitch, or energy of the signal, although these categories often overlap (**Table 2**). An example of a time-related feature is the pause rate, which measures how often a speaker takes pauses of a prescribed length during a time period.^24^ An informative pitch-related feature is the Mel-frequency cepstrum coefficient (MFCC), which rescales frequencies of the audio signal logarithmically to mimic human auditory perception.^25^ Finally, an energy-related feature is the loudness of a sound, typically measured in decibels.^25^ Several tools are available for feature extraction from audio signals (**Table 1**), of which, the open-source Praat^26^ and OpenSMILE^27^ the most widely used.

**Table 2:** Features commonly used in audio analysis.

| Category | Feature | Definition |
| --- | --- | --- |
| Time | Pause rate<sup>24</sup> | Rate (instances per unit of time) at which an audio pauses for a duration exceeding a predefined threshold. |
| (features mostly related to how the signal changes over time) | Phonation rate<sup>24</sup> | Total phonation time (duration of real signal) divided by total duration of the audio. |
|  | Fundamental frequency trajectory<sup>25</sup> | Changes in the fundamental frequency (lowest frequency) emitted over the audio |
| Pitch | Jitter<sup>25</sup> | Changes in the length of consecutive occurrences of the fundamental frequency |
| (features mostly related to the frequency content of the signal) | Formants ($F_1$, $F_2$, ...) <sup>99</sup> | Frequencies of resonances caused by the shape of the vocal tract, typically during the phonation of a vowel<sup>16</sup> |
|  | Mel-frequency cepstrum coefficients (MFCC) <sup>25</sup> | Coefficients derived from filtering and scaling frequencies according to how human hearing typically perceives them |
| Energy | Loudness<sup>25</sup> | Perceived intensity of an acoustic signal |
| (features mostly related to the energy of the signal) | Harmonic Difference<sup>25</sup> | Ratio of energies contained in different pairs of harmonics*, often consecutive |
|  | Harmonic to Noise Ratio (HNR) <sup>25</sup> | Ratio of harmonic components of a signal to the underlying signal noise |
| * naturally occurring frequencies at integer multiples of a fundamental frequency<sup>16</sup> |  |  |

It can often be unfavorable for the computational efficiency and performance of data analysis methods to use too many features due to the *curse of dimensionality* problem.^28^ Thus, the Geneva Minimalist Acoustic Parameter Set (GeMAPS) has been proposed as a minimalist set of 62 features that are expected to be effective for audio.^25^ GeMAPS is widely used due to its inclusion in openSMILE.^27^

### Prediction methods for clinical audio

Automated audio processing methods have been widely used for predicting disease indications, especially in neuropsychiatry. We describe these efforts in the following subsections.

### Statistical and traditional machine learning (ML) methods

Statistical methods, such as hypothesis testing, have been used routinely to analyze biomedical data.^29^ Traditional ML methods, e.g., Support Vector Machine (SVM), Random Forest (RF) and k-Nearest Neighbor (kNN), have also been widely applied in biomedicine.^11^ These methods are designed to sift through large amounts of data, structured or unstructured, without any particular guiding (biomedical) hypothesis, to discover potentially actionable knowledge. A *predictive model* encapsulates a mathematical relationship between the data describing an entity of interest, say a patient, and an outcome or label, say the disease status, of the entity. The purpose of this model is to make predictions of this outcome or label that are not yet known for other entities.

Early applications of these methods aimed to understand which features of clinical audio had explanatory or predictive power.^30–33^ A semi-automated approach assessed speech differences between children with cerebral palsy and controls by analyzing data from speech elicitation tasks.^30^ Trained listeners transcribed speech recordings, which were used to determine word counts and what proportion of the words uttered matched the target elicitation (intelligibility). The study found that speech rate (words uttered per minute) and intelligibility classified normally developing children and those with cerebral palsy.

A similar study analyzed speech from picture-describing and sentence-repeating tasks to distinguish between patients with Alzheimer’s dementia (AD) and those with mild cognitive impairment (MCI).^31^ The study found that the duration of speech and the increased likelihood of inserting or deleting to words in prompted sentences differentiated AD and MCI patients. Other studies used statistical tests like Mann-Whitney U to evaluate the association of pitch features (**Table 2**) with neuropsychiatric conditions.^31–33^

Several studies also used audio features and ML methods (**Table 2**) to classify patients into classes corresponding to neuropsychiatric conditions.^31,34–36^ Most of these studies use variations of the SVM algorithm, which finds an optimal boundary separating two classes of data points. One study used multiple SVM models to identify a motor speech disorder by determining the severity of unintelligible speech.^35^ A sentence-level SVM trained on energy features and a phoneme-level SVM trained on pitch and time features yielded accuracies of 79.8% and 77.3% respectively. This accuracy increased to 84.8% when the SVMs were combined, an approach known as ensemble learning.^11^ Other studies have found success in similar tasks using other traditional ML algorithms like RF and kNN.^37,38^

Several approaches also combined audio features with linguistic characteristics derived from textual transcripts of the audio.^32,36,38,39^ One study performed 3-way classification of levels of cognitive impairment (control, mild, and early stage Alzhiemer’s disease) using an SVM trained on acoustic features derived using Praat^26^ and an automatic speech recognition (ASR) system (**Table 1**).^40^ This classifier had an accuracy of 60%, which improved to 66.7% after combining the acoustic features with a variety of linguistic ones.

### Deep learning

More recently, deep learning (DL) techniques^13^ have been used to characterize clinical conditions from patient audio.^10^ These techniques typically utilize much larger, but not always explicitly specified, feature sets than statistical and traditional ML techniques. DL techniques generally utilize multi-layer neural networks to build implicit representations of datasets and enable various analysis tasks, including predictive modeling (**Figure 3**).^12^ Due to this architecture, DL techniques are capable of building predictive models directly from native audio recordings.

**Figure 3:**
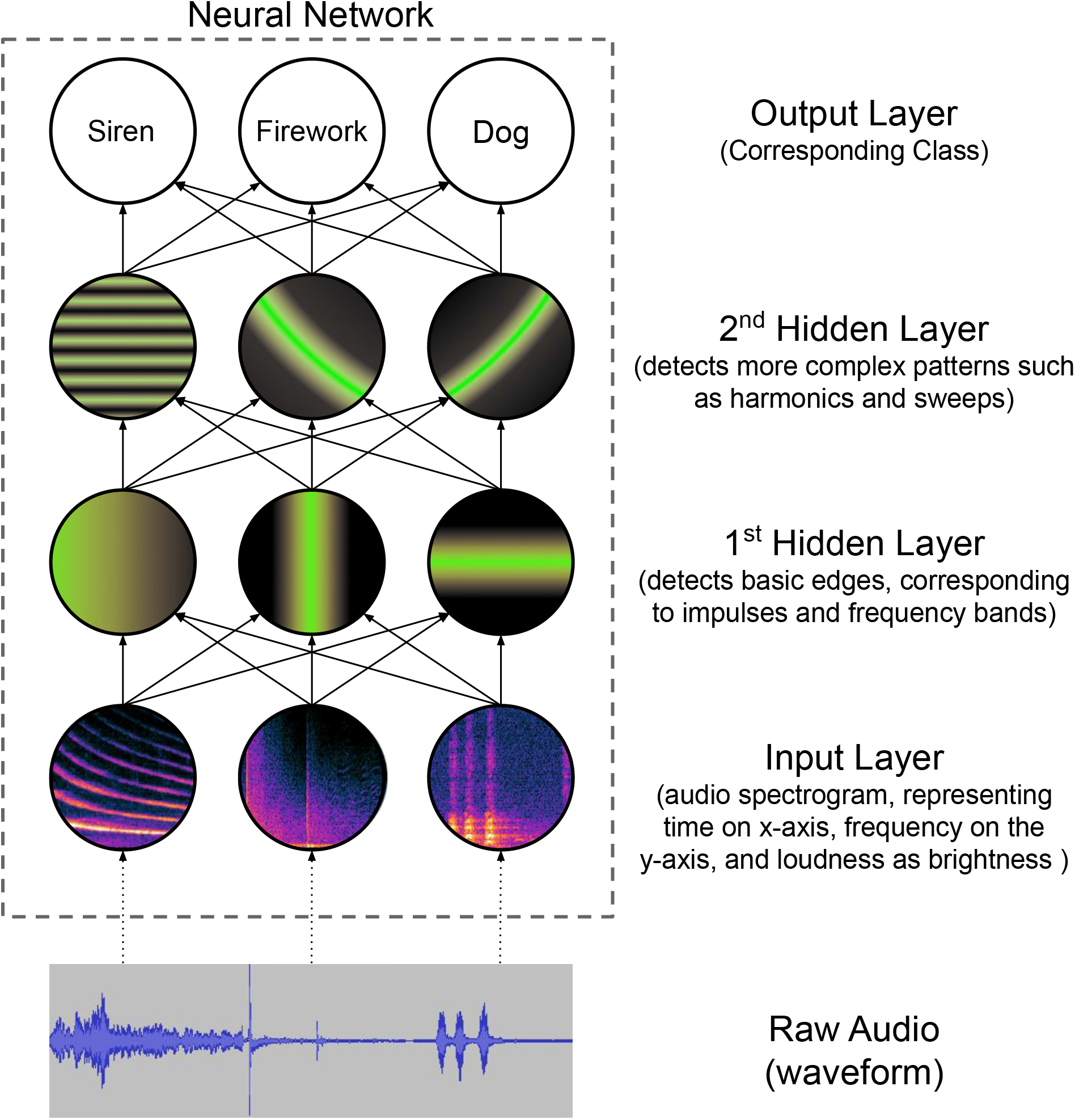
An illustration of a deep learning-based neural network model that can be used to represent audio at various levels of abstraction. The raw audio is input to the network at the waveform level, and at each successive “layer” of the network, a more abstract representation is learned. At the final layer of the network, the model predicts the most likely class that the audio originated from.

Convolutional neural networks (CNNs) (**Figure 4**) are among the most prevalent DL techniques for audio and other unstructured data, especially due to their ability to represent contextual information in data.^13^ Another established architecture is the recurrent neural network (RNN) (**Figure 5**), where sequentially structured inputs pass through functional transformations at consecutively connected layers of the network.^13^

**Figure 4:**
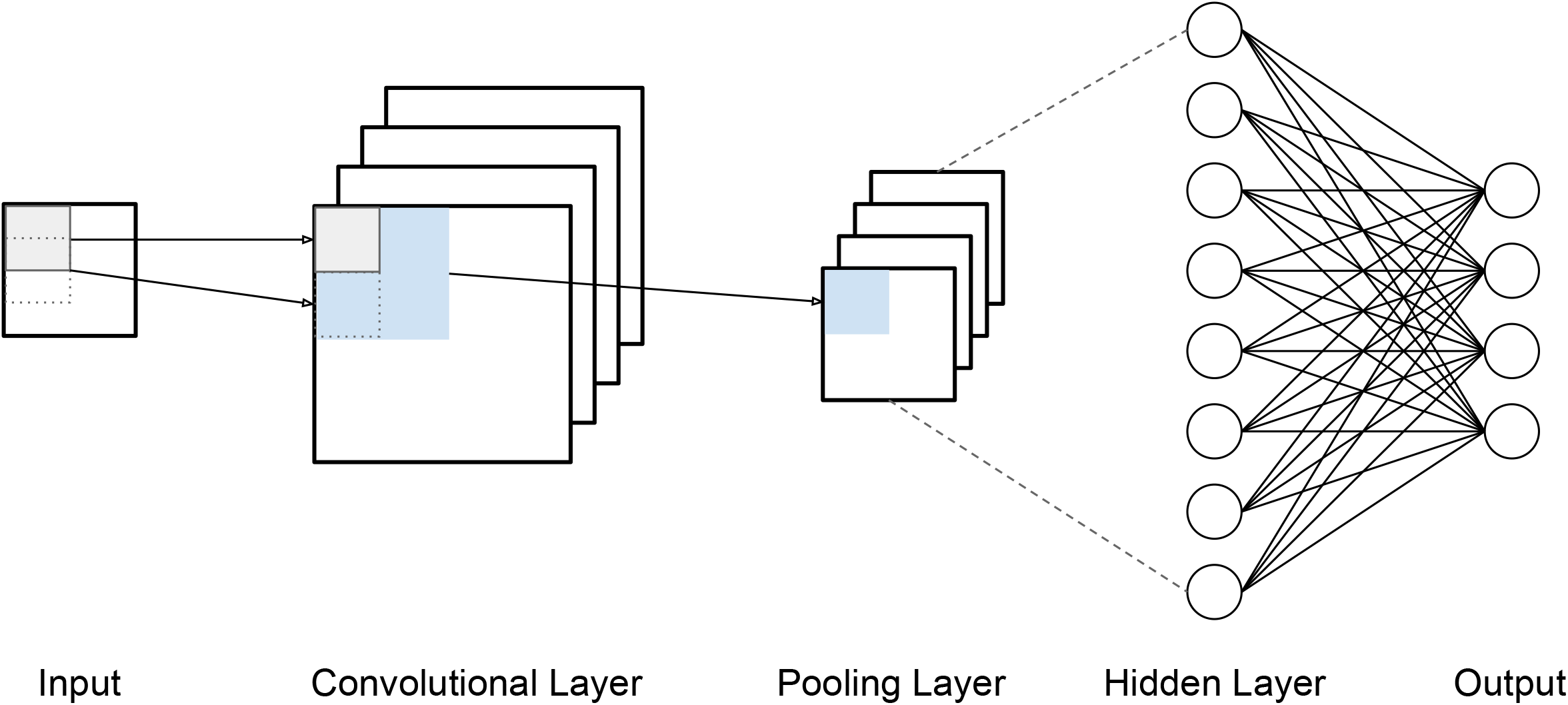
The architecture of a Convolutional Neural Network (CNN). Input data are passed through a convolutional filter to extract important contextual information embedded in the data through a mathematical function. The intuition behind convolutions is to mimic human neuronal processes, wherein certain neurons attend to specific parts of sensory stimuli.^12^ These convolved inputs are then pooled to reduce dimensionality, and eventually passed through fully connected layers to generate classifier outputs.

**Figure 5:**
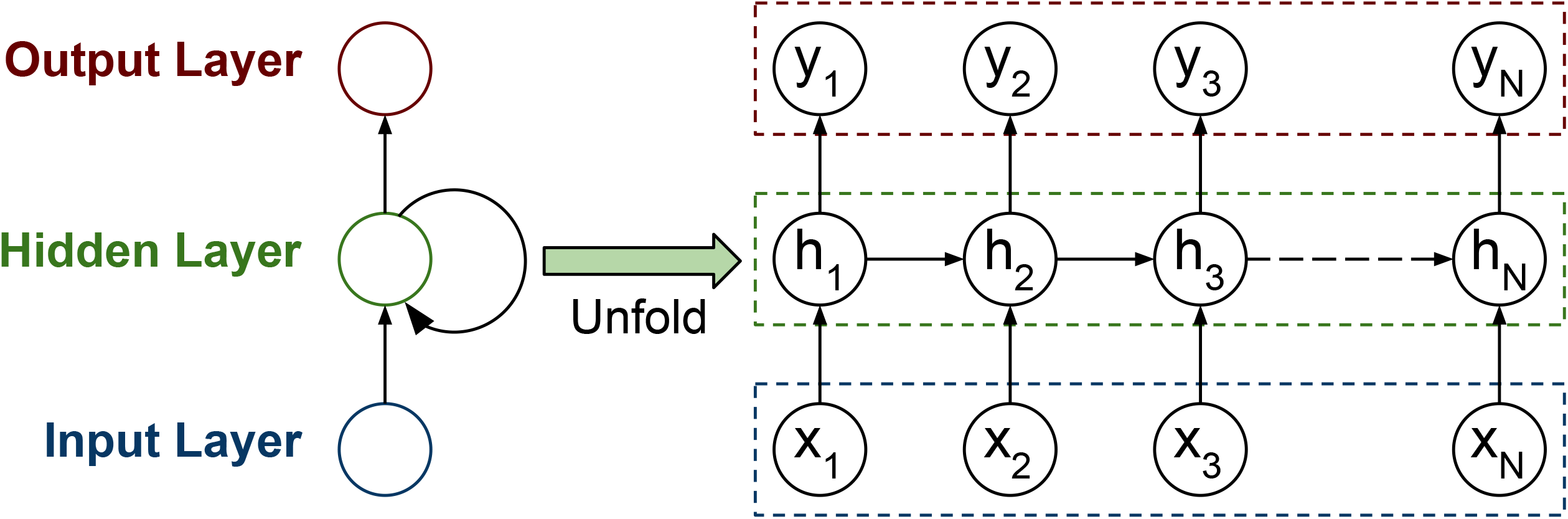
The architecture of a Recurrent Neural Network (RNN). Ordered inputs are fed sequentially into a neural network. The “unfolding” of the network is designed to infer hidden states h_n_, which are a function of the current input x_n_ and of the previous hidden state h_n-1_, thus capturing the “recurrent” nature of the network. The learned representation embedded in these states is then used to produce the desired output at different points in time or in the sequence. This sequential structure of RNNs makes them particularly suitable for time series data like language, audio, and speech.

Most clinical audio processing that utilizes DL inputs pre-computed features into a CNN, which can then predict the presence or degree of a condition.^41,42^ One investigation built a CNN model using GeMAPS^25^ and other feature sets to classify depression severity.^43^ A related approach trained parallel CNN models for the different categories of audio features in **Table 2**.^43^ The last dense layers of these CNNs were then concatenated to predict depression severity. Multiple investigations found that an ensemble of individual CNNs built from different data modalities (e.g., audio, text, and video) can predict depression severity even more accurately.^41,42,44^

Other approaches have leveraged the sequential or temporal nature of audio. One AD detection effort employed a Time-Delayed CNN.^45^ Instead of the entire recording, this approach applied the convolutional filter to utterances (segments of speech separated by silence) over all preceding time frames (hence the “delay”). This allowed them to extract local features from different temporal segments of the recording.^45^ Another study used a Long-Short Term Memory architecture (LSTM, a sophisticated implementation of an RNN)^12^ to screen for depression.^46^ MFCCs (**Table 2**) were extracted from different temporal segments of the audio and input to the LSTM. The outputs of the recurrent layers were then fed to the fully connected layers of the network to predict depression scores.^46^

DL methods have also been used in situations of insufficient audio data. One study^46^ employed transfer learning^12^ to classify eight types of emotions from the relatively small RAVDESS dataset^47^ (**Table 3**). This approach repurposed an RNN trained on a data-rich task (depression score classification), and fine-tuned it with RAVDESS for a related task (emotion recognition). The approach achieved a validation accuracy of 76.3%, an increase of 8.7% from a baseline RNN trained solely on the RAVDESS.^46^ Another useful DL architecture for data augmentation is a Generative Adversarial Network (GAN),^12^ which consists of two competing neural networks, a generator and a discriminator, to generate new data samples. In an approach to diagnose childhood autism,^48^ the generator of a GAN was trained using GeMAPS^25^ and other feature sets extracted from the Child Pathological Speech Database^34^ (**Table 3**), and a discriminator was trained to help the model generate more realistic data points. Learned representations of the data were then extracted from the intermediate layers of the discriminator, and used to train an SVM model to classify four levels of pathology related to developmental disorders and autism.

**Table 3:**
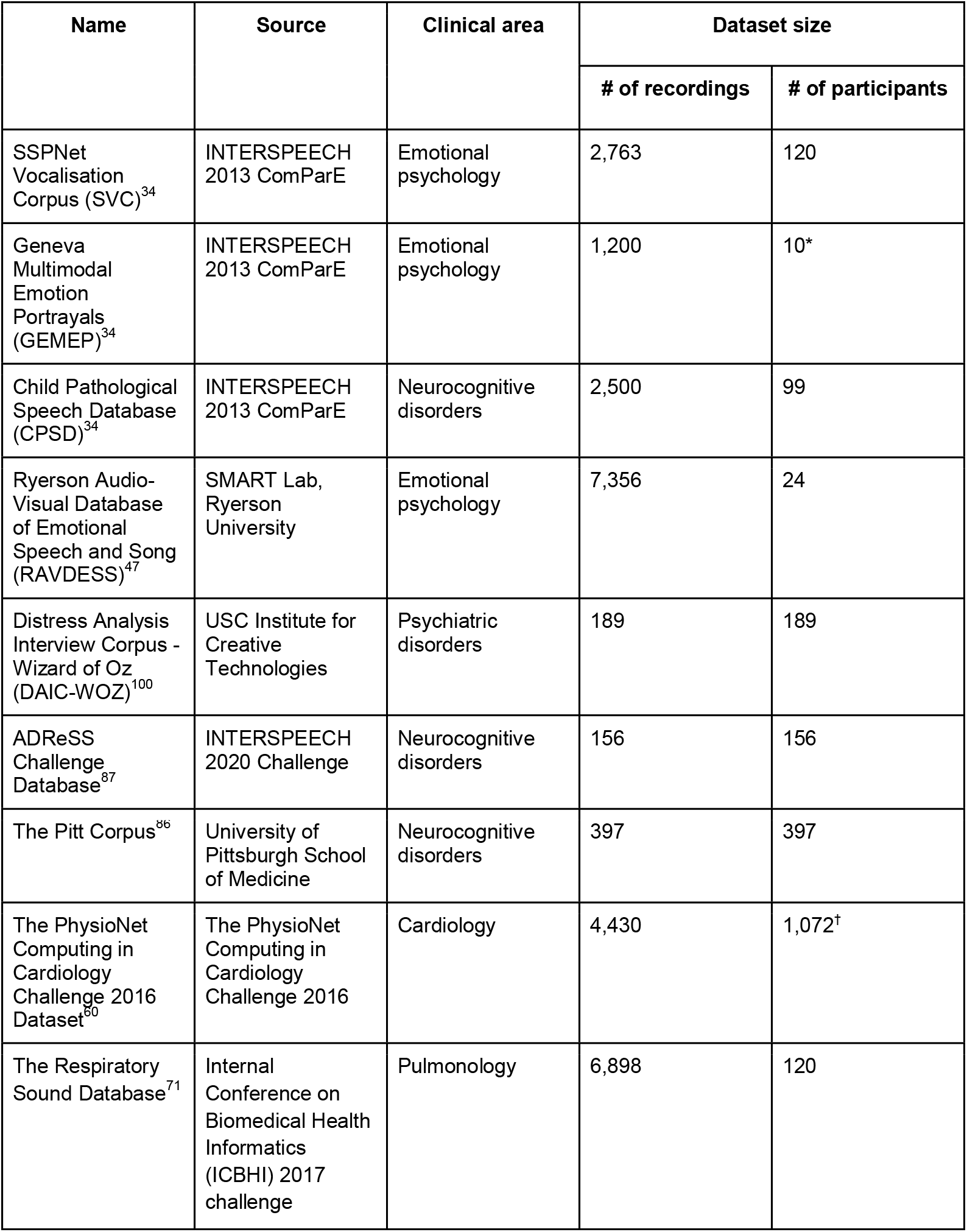
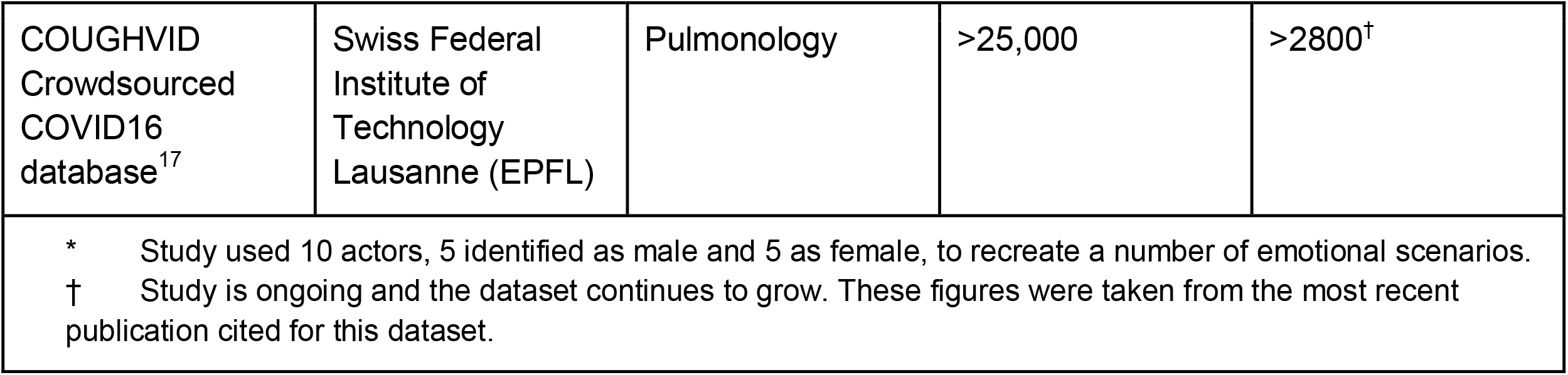
Details of prominent crowdsourced challenges and publicly available datasets.

| Name | Source | Clinical area | Dataset size |  |
| --- | --- | --- | --- | --- |
|  |  |  | # of recordings | # of participants |
| SSPNet Vocalisation Corpus (SVC)<sup>34</sup> | INTERSPEECH 2013 ComParE | Emotional psychology | 2,763 | 120 |
| Geneva Multimodal Emotion Portrayals (GEMEP)<sup>34</sup> | INTERSPEECH 2013 ComParE | Emotional psychology | 1,200 | 10* |
| Child Pathological Speech Database (CPSD)<sup>34</sup> | INTERSPEECH 2013 ComParE | Neurocognitive disorders | 2,500 | 99 |
| Ryerson Audio-Visual Database of Emotional Speech and Song (RAVDESS)<sup>47</sup> | SMART Lab, Ryerson University | Emotional psychology | 7,356 | 24 |
| Distress Analysis Interview Corpus - Wizard of Oz (DAIC-WOZ)<sup>100</sup> | USC Institute for Creative Technologies | Psychiatric disorders | 189 | 189 |
| ADReSS Challenge Database<sup>87</sup> | INTERSPEECH 2020 Challenge | Neurocognitive disorders | 156 | 156 |
| The Pitt Corpus<sup>86</sup> | University of Pittsburgh School of Medicine | Neurocognitive disorders | 397 | 397 |
| The PhysioNet Computing in Cardiology Challenge 2016 Dataset<sup>80</sup> | The PhysioNet Computing in Cardiology Challenge 2016 | Cardiology | 4,430 | 1,072<sup>†</sup> |
| The Respiratory Sound Database<sup>71</sup> | Internal Conference on Biomedical Health Informatics (ICBHI) 2017 challenge | Pulmonology | 6,898 | 120 |
| COUGHVID<br>Crowdsourced<br>COVID16<br>database<sup>17</sup> | Swiss Federal<br>Institute of<br>Technology<br>Lausanne (EPFL) | Pulmonology | >25,000 | >2800<sup>†</sup> |
| * Study used 10 actors, 5 identified as male and 5 as female, to recreate a number of emotional scenarios.<br>† Study is ongoing and the dataset continues to grow. These figures were taken from the most recent publication cited for this dataset. |  |  |  |  |

Some approaches use *x-vectors*, which are DL-based representations trained for speaker identification in a conversation.^49,50^ These *x-vectors* can then be used with ML or DL methods to classify speakers with and without a pathology.^51^ Several studies reported better performance of this approach for Alzheimer’s and Parkinson’s disease diagnosis compared to feature-based methods.^51–54^ Another study utilized an alternative Active Data Representation (ADR).^55^ built for speakers from the Pitt Corpus (**Table 3**), and used them to classify AD, performing better than traditional ML methods.

Finally, recent DL methods learn which characteristics of a native audio signal are useful for classification. Zhao and colleagues used a hierarchical attention transfer network that reconstructed segments of patient speech using an autoencoder, and integrated them with LSTM representations from a speech recognition model to screen for depression.^56^ Another study compared using the raw audio signal, and its various filtered versions, with a CNN.^57^ Filtering the signal boosted the CNN’s ability to accurately classify levels of depression from patient speech, illustrating the advantages of effective pre-processing.

## Other clinical applications

While most of the research in this area has been on cognitive health, similar work is emerging in other realms, as described below.

### Cardiac conditions

High-fidelity recordings of heart sounds can be collected using digital stethoscopes or phonocardiograms (PCGs).^58^ After pre-processing to remove ambient noise, the recordings are generally segmented to isolate each beat and its components, traditionally using thresholds of acoustic features defined by clinical experts, or Gaussian probabilistic models.^59^ A recent study used a Hidden Markov Model (HMM) to segment PCG recordings into four stages of a heartbeat, namely S1, systole, S2, and diastole.^59^

Several studies have also developed classifiers of cardiovascular disease from segmented PCGs. The 2016 PhysioNet Computing in Cardiology Challenge (**Table 3**) produced several accurate classifiers for these diseases.^11,60^ The best-performing method used an ensemble of a feature-based adaptive boosting model and a CNN.^61^ The runner-up used an ensemble of 20 neural networks.^62^ More recent studies have found similar success using DL techniques.^63,64^

### Respiratory diseases

The most common use of audio for respiratory diseases has been the identification and classification of coughs. Several studies classified coughs, most commonly as “wet” or “dry”, to help distinguish lower respiratory tract infections like pneumonia, bronchitis, and tuberculosis from other diseases.^65,66^ These studies used features extracted from audio (**Table 2**) and classifiers like logistic regression.

Similar traditional ML and some DL methods have been used to directly classify specific respiratory diseases, such as pneumonia, asthma, and whooping cough.^65–70^ A representative study compared traditional ML and DL methods for this task using the Respiratory Sound Database (**Table 3**)^71^, and found that the best-performing models were CNN, LSTM, and an ensemble of CNN and RNN.

Finally, a number of studies have attempted to diagnose COVID-19 from recordings of patients’ coughs.^72^ Researchers crowdsourced the COUGHVID dataset (**Table 3**) of cough recordings using a web-app accessible from mobile devices, a subset of which were labeled by pulmonologists in terms of the type of cough and other characteristics.^17^ The same effort proposed an algorithm based on XGBoost^73^ to filter out recordings containing no coughs. DL methods using extracted audio features with CNN, LSTM, and ResNet architectures have found the most success for classifying coughs and diagnosing COVID-19.^74,75^

### Sleep disorders

Studies on sleep disorders attempted to quantify snoring and identify conditions like obstructive sleep apnea and sleep-disordered breathing from data typically recorded using bedside^76^ or overhead^76^ microphones and smartphones^77^ in a controlled environment. Several of these studies employed DL methods with some success, although the quality and quantity of available data remains a problem.^54,78^

### Other Applications

Audio processing and ML continue to be applied to a variety of new clinical problems, such as automatically detecting seizures,^79^ improving the quality of hearing aids,^80^ and identifying out-of-hospital cardiac arrests from emergency dispatchers.^81^

## Evaluations methodologies, benchmarking and open issues

### Evaluation methodologies

As in general ML, rigorously evaluating audio-based predictive models is as important as building them. Several of the studies reviewed above evaluated these models on a separate test and validation set. If a test set was not available, studies often used bootstrapping or cross-validation,^11^ which repeatedly use different randomly chosen partitions of the training set to develop and evaluate the models.

Several evaluation measures have been proposed to quantify the performance of classification-oriented predictive models (**Figure 6**).^82^ Accuracy and the area under the Receiver Operating Characteristic (ROC) curve (AUC score) were the most routinely used evaluation measures. However, these measures can be misleading in cases where the classes are imbalanced, which is common in biomedical sciences.^82,83^ In these cases, it is recommended to use the class-specific Precision, Recall and F-measure metrics (**Figure 6**) for evaluation.

**Figure 6:**
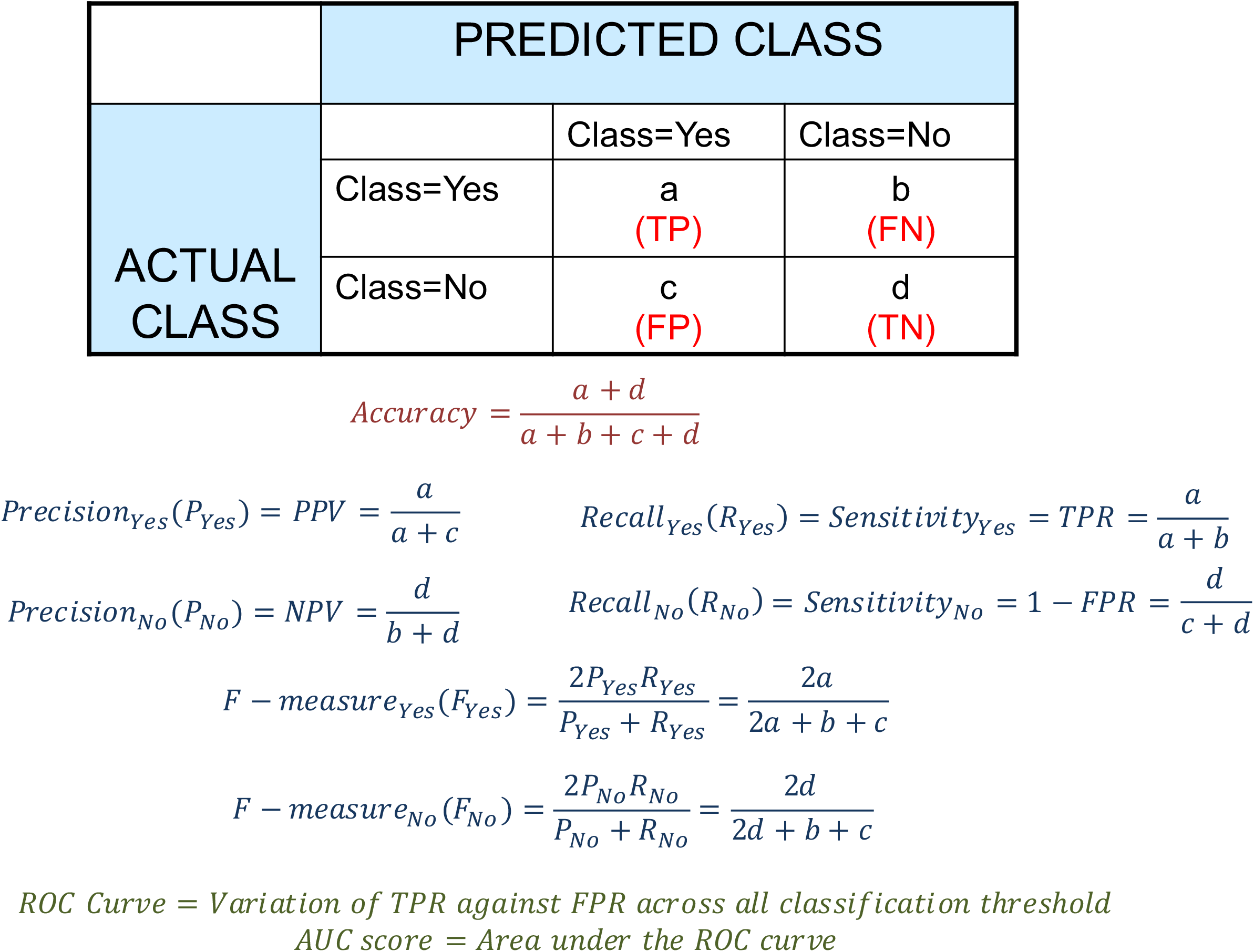
A binary confusion matrix for classification models that can be used to evaluate several evaluation measures. The definition of and relationships between these measures are summarized. This matrix and these measures can be similarly defined for more than two classes.

### Benchmarking using crowdsourced challenges and public datasets

It can be difficult to gauge the general effectiveness of a new ML tool for audio analysis unless it is evaluated against an established benchmark dataset.^84^ Since it is laborious for individual studies or laboratories to collect large datasets, *crowdsourced challenges* have been organized to address this important need.^85^ **Table 3** provides the details of several prominent challenges and publicly available datasets.

### Issues facing the field

Although substantial progress has been made in automated analysis of clinical audio, the field still faces several interrelated issues that should be addressed to enable further progress.

### Insufficient data

Often, the most significant problem for any data-driven study is insufficient data. In particular, recording clinical audio introduces a number of challenges. These include the cost of a recording device, distraction from providing care, and the need for patient consent. As a result, several studies reviewed above relied on relatively small datasets. This issue is particularly challenging for DL, which typically requires large training datasets.^13^ Although benchmark datasets (**Table 3**) and data augmentation^12,20^ methods can address this problem to some extent, a general paucity of sufficiently large datasets in multiple clinical areas poses a significant issue for the area.

### Incompatibility of data from multiple sources

The problem of insufficient data is partially caused by the incompatibility of audio collected from multiple sources. Even in studies on the same disease, the respective datasets may be incompatible due to differing recording protocols and methodologies. For instance, both the Pitt Corpus^86^ and the 2020 ADReSS Challenge dataset^87^ (**Table 3**) include recordings of dementia patients and controls. However, due to different questionnaires, recording hardware, and pre-processing, the two datasets are quite incompatible. As a result, a model trained on one is unlikely to perform well on the other. The absence of a standard approach to data collection in the field hinders progress.

### Reproducibility of features

An issue with audio feature-based approaches is that the features themselves are not always reproducible. Some studies found that even features extracted with established software (**Table 1**) do not produce sufficiently consistent results when compared across recordings.^26,27^ While robust ML and DL approaches are able to address this problem to some extent by focusing on the important information in the features, it is difficult to complete resolve this data reliability problem algorithmically.

### Biases and confounding factors

As in many applications of ML, the performance of audio analysis tools may be limited by the characteristics of their training data, raising concerns about bias, diversity and inclusion. For instance, the vast majority of the work in the field is conducted with recordings of English-speaking individuals, raising questions about their effectiveness for other languages. Some tools may also be affected by confounding factors like dialect, accent, age, ethnocultural status (e.g., race/ethnicity), gender and medication regimen.^88,89^ Few studies reviewed here reported participants’ demographic characteristics, further exacerbating this problem. Future studies should use as inclusive and representative datasets and algorithms as possible, and consider confounding factors.

## Conclusion

This review described a broad spectrum of work in automated audio processing that aimed to accurately characterize the clinical status of patients. These studies utilized as large datasets of audio as possible, and a wide array of statistical, traditional ML and DL techniques. Most of this work focused on neuropsychological conditions, but interesting work in other clinical areas (e.g., cardiac, infectious and sleep disorders) is emerging. The review also discussed several important challenges facing the field, as well as potential ways to address them.

The ultimate goal of this area is to clinically operationalize these algorithms to aid in monitoring and diagnosing diverse conditions. However, most of the efforts to date have been in laboratory settings, focusing on refining analytic processes. While there has been some movement towards standardization and benchmarking, there has been limited deployment of these methods. Some software systems that can be shared among clinicians have been developed. NeuroSpeech is one such system that can help analyze dysarthric speech and diagnose Parkinson’s disease.^90^ Another group deployed a smartphone application to recognize phonemic boundaries and interpret a patient’s speaking rate for real-time speech therapy.^91^

Future work in this direction will have to address several challenges. First, an infrastructure for the recording, processing and storage of high-quality patient audio will have to be established and standardized.^8,11^ These data must be obtained with patients’ consent, and be stored securely to maintain privacy and confidentiality.^13^ Finally, institutions may also need high-performance computing facilities for this deployment, especially to develop predictive models for their patient populations.^92^ As progress is made in these directions, we expect that audio-based tools will be deployed in the clinic much more extensively in the future.

## Data Availability

Not applicable.

## Acknowledgements

This work was supported by NIH grant #s R01 AG066471 and R01 HG011407-01A1.

## Author Contributions

A.K., A.F., G.P., and T.J. conceived this review. A.K. and T.J. analyzed the literature, collected the data and drafted the manuscript under G.P.’s supervision. All the authors contributed to the drafting of the manuscript and approved the final version.

